# Comprehensive profiling of social mixing patterns in resource poor countries: a mixed methods research protocol

**DOI:** 10.1101/2023.12.05.23299472

**Authors:** Obianuju Genevieve Aguolu, Moses Chapa Kiti, Kristin Nelson, Carol Y. Liu, Maria Sundaram, Sergio Gramacho, Samuel Jenness, Alessia Melegaro, Charfudin Sacoor, Azucena Bardaji, Ivalda Macicame, Americo Jose, Nilzio Cavele, Felizarda Amosse, Migdalia Uamba, Edgar Jamisse, Corssino Tchavana, Herberth Giovanni Maldonado Briones, Claudia Jarquín, María Ajsivinac, Lauren Pischel, Noureen Ahmed, Venkata Raghava Mohan, Rajan Srinivasan, Prasanna Samuel, Gifta John, Kye Ellington, Orvalho Augusto Joaquim, Alana Zelaya, Sara Kim, Holin Chen, Momin Kazi, Fauzia Malik, Inci Yildirim, Benjamin Lopman, Saad B. Omer

## Abstract

**Background:** Low-and-middle-income countries (LMICs) bear a disproportionate burden of communicable diseases. Social interaction data inform infectious disease models and disease prevention strategies. The variations in demographics and contact patterns across ages, cultures, and locations significantly impact infectious disease dynamics and pathogen transmission. LMICs lack sufficient social interaction data for infectious disease modeling.

**Methods:** To address this gap, we will collect qualitative and quantitative data from eight study sites (encompassing both rural and urban settings) across Guatemala, India, Pakistan, and Mozambique. We will conduct focus group discussions and cognitive interviews to assess the feasibility and acceptability of our data collection tools at each site. Thematic and rapid analyses will help to identify key themes and categories through coding, guiding the design of quantitative data collection tools (enrollment survey, contact diaries, exit survey, and wearable proximity sensors) and the implementation of study procedures.

We will create three age-specific contact matrices (physical, nonphysical, and both) at each study site using data from standardized contact diaries to characterize the patterns of social mixing. Regression analysis will be conducted to identify key drivers of contacts. We will comprehensively profile the frequency, duration, and intensity of infants’ interactions with household members using high resolution data from the proximity sensors and calculating infants’ proximity score (fraction of time spent by each household member in proximity with the infant, over the total infant contact time) for each household member.

**Discussion:** Our qualitative data yielded insights into the perceptions and acceptability of contact diaries and wearable proximity sensors for collecting social mixing data in LMICs. The quantitative data will allow a more accurate representation of human interactions that lead to the transmission of pathogens through close contact in LMICs. Our findings will provide more appropriate social mixing data for parameterizing mathematical models of LMIC populations. Our study tools could be adapted for other studies.

## Introduction

Infectious diseases account for almost half of all child deaths worldwide.(1) Diarrhea, lower respiratory infections, and malaria are the leading causes of death among the 5-14-year-old age group.(1) The world’s poorest billion bear a disproportionate burden of these diseases.(2) For example, 91% of all deaths due to diarrheal diseases and 64% of all deaths due to respiratory illnesses occur in low-and-middle-income countries (LMICs).(3) The recent increase in the incidence of emerging infectious diseases and the recent COVID-19 pandemic have placed extreme stress on already fragile healthcare systems.(4,5) Global structural inequalities further exacerbate COVID-19-related health disparities in the LMICs, such as delayed access to vaccines and therapeutics.(6) During earlier phases of the pandemic, nonpharmacological strategies such as quarantine, physical distancing, stay-at-home orders, and work-from-home policies were the major tools used globally to mitigate the spread of the virus.(7) Mathematical models of infectious diseases based on surveillance data can be used to address both scientific hypotheses and infectious disease-control policy questions and are increasingly influential in informing health policy and investment strategies globally.(8)

Direct data on social contacts collected for eight high-income European populations in the POLYMOD study between May 2005 and September 2006 have informed many models with resulting policies.(9–20) However, demographic structures and contact data differ between populations. The structure of human contact networks varies by age, culture, and location with profound implications for pathogen dynamics and transmission.(20) Social contact patterns may differ in different geographic areas due to a variety of differences in social institutions, population density and family structure.(20–22) Despite these differences, there is a lack of systematically collected data on social mixing in LMICs. Models specifically parameterized to LMICs that incorporate localized social mixing patterns are essential to inform public health policies.

While some studies have collected social mixing data in low-income settings,(11–20) they were conducted in single populations using varying approaches. Therefore, it is difficult to infer whether differences represent true variation between populations or are an artifact of study design. Furthermore, data have traditionally been collected using paper-based contact diaries, which are subject to recall errors and bias. Recently, automated methods of data collection utilizing portable technology such as mobile phones and low-powered Bluetooth wearable sensors have become popular.(23) Thus, social mixing study data are limited for parameterizing models of infectious diseases in LMICs. Information on changes in contact patterns during times when physical distancing measures were implemented (for example, as part of a public health response to the COVID-19 pandemic) can further inform mathematical models, by noting the maximum reduction in social contacts that is realistically achievable in restrictive environments.(24)

We propose a multicountry mixed methods (qualitative and quantitative) study with the overall goal of using standardized methods (contact diaries and wearable proximity sensors) to collect social contact data from urban and rural populations in LMICs. The contact diary will be used to collect self-reported data on participants and their contact(s), as well as individual characteristics of the person they had contact with, such as age, sex, and relationship to the contact person. The wearable proximity sensor is powered by a battery and uses a low-power active-radio frequency identification (RFID) technology to transmit and receive signals when hung around the neck or worn in a shirt/blouse pocket. This device will be used to collect contact data among family members of infants younger than 6 months old. The study will include individuals from all age groups. Furthermore, special focus will be given to studying the social interactions of infants younger than six months of age. Data will be rigorously collected from four different LMICs to capture a diverse cross-section of geographic locations: Guatemala (Central America), India and Pakistan (South Asia) and Mozambique (Sub Saharan Africa). These sites were selected due to the presence of a strong community-based infrastructure, existing disease and demographic surveillance systems, and history of collaborating with the study team, and to ensure geographic diversity. The data collected in this study will be made publicly available to be used in transmission dynamic models aiming to accurately represent human interactions that lead to transmission of respiratory and gastrointestinal infections.

## Materials and Methods

This is a multicountry cross-sectional survey. Our main study objective is to characterize social contact patterns in urban and rural settings in low-and-middle-income countries. Specifically, we aim to i) Explore the perceptions and acceptability of paper contact diaries and wearable proximity sensors to collect data on social contact and mixing patterns between individuals; ii) Characterize the patterns of social contact and mixing in urban and rural settings using standardized social contact diaries; and iii) Comprehensively profile the social contacts of infants with their household members using wearable proximity sensors. We will use qualitative (formative phase) and quantitative methods for our data collection and analysis. Data will be collected in four countries: Mozambique, Guatemala, India, and Pakistan (table 1, figure 1), one rural and one urban site, for a total of 8 study sites in each country.

**Table 1.** Study site characteristics

| Country | Mozambique |  | Guatemala |  | India |  | Pakistan |  |
| --- | --- | --- | --- | --- | --- | --- | --- | --- |
| Study site | Rural | Urban | Rural | Urban | Rural | Urban | Rural | Urban |
|  | Manhiça District, Maputo Province | Bairro Polana Caniço, Maputo City | Concepción Chiquirichapa: Telená, Tuilcanabaj (Centro/Bodega) San Juan Ostuncalco: Monrovia and La Esperanza in the Quezaltenango Department | San Juan Ostuncalco: Centro San Juan. Concepción Chiquirichapa: Centro Concepción | Kilarasampet, Kathalampet in Kaniyambadi Block, Vellore district, Tamil Nadu. | Ward 59, Vellore corporation | Matiari | Ali Akber Shah Goth in Karachi East |
| Source of participant data | Existing demographic surveillance system (DSS) in Manhiça and Polana Caniço |  | Randomized sampling from selected households in 57 clusters identified by aerial imaging |  | Existing DSS |  | Existing DSS for <5-year-old and 15–49-year-old women in Karachi; |  |

**Figure 1.**
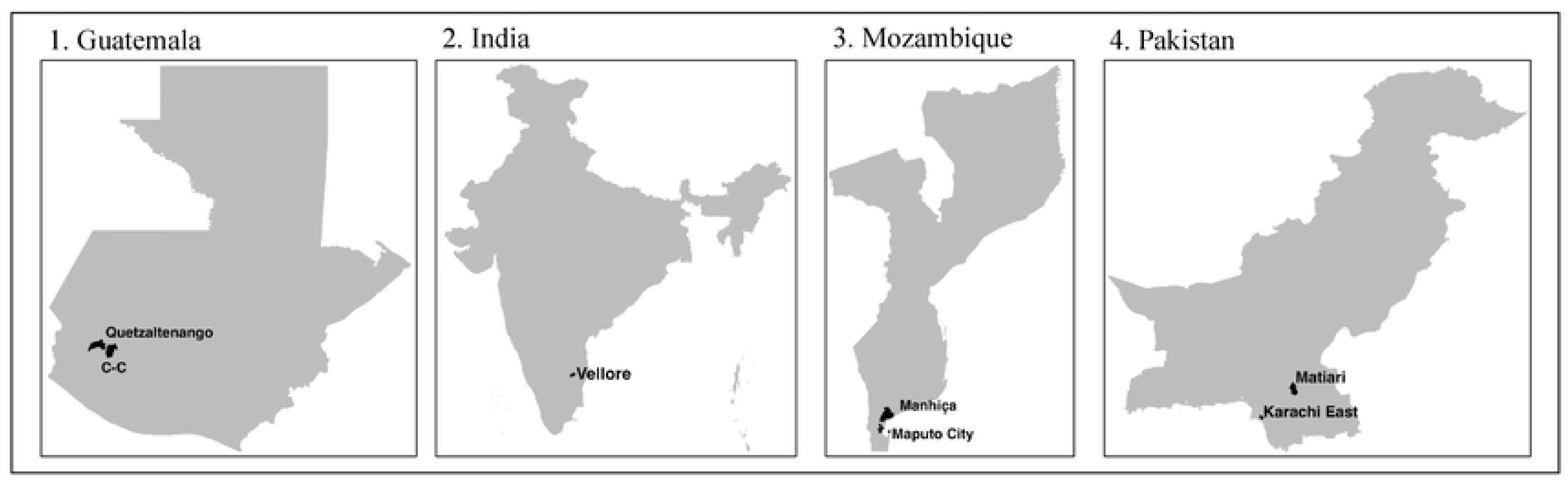
Global map showing the location of Guatemala, India, Mozambique, and Pakistan. Each country has one rural and one urban site. There are separate rural (Manhica and Matiari) and urban (Maputo and Karachi East) sites in Mozambique and Pakistan, respectively. In Guatemala, rural and urban clusters will be identified from the Quetzaltenango and xxx. In India, rural and urban areas will be selected from Vellore.

## Data collection tools

For the qualitative phase, data were collected through focus group discussions (FGDs) and psychometric testing through cognitive interviews. For the quantitative phase, data will be collected using an enrollment survey, paper and electronic contact diaries, an exit survey, and wearable proximity sensors.

### Enrollment survey

The enrollment survey will be used to enroll participants after they provided consent. It contains questions on demographic details of the participant including age, sex, school enrollment, occupation, study site, as well as household characteristics such as household size and composition, and sources of water.

### Contact diary

We developed paper and electronic contact diaries to collect data on participants’ social mixing. We defined contact as:

- Nonphysical contact: This involves a two-way conversation with three or more words exchanged in the physical presence of another person. ·
- Physical contact: This involves directly touching someone (skin-to-skin contact) or the clothes they are wearing (for example a handshake, fist bump, elbow bump, foot bump, hug, kiss, etc.).
- Direct proximity: This involves being within 6 feet for 20 seconds or more.

The paper diary will be given to participants to self-report data on their contacts and characteristics of the person with whom they had contact, such as age (approximate where appropriate), sex, relationship to the contact person, location, frequency, and duration of contact. The electronic contact diary was designed in REDCap and downloaded onto mobile tablets used by the field staff. For each site, we developed a set of culturally relevant images representative of the 10 age groups that a contact may fall into. We classified the age groups as follows: <6 months, 6-11 months, 1-4 years, 5-9 years, 10-14 years, 15-19 years, 20-29 years, 30-39 years, 40-59 years, and 60+ years. Participants will use the set of images to give estimates of the age of their contacts. Each tool was translated to the major local languages for each study site (Portuguese in Mozambique; Spanish in Guatemala; Tamil in India, and Urdu and Sindhi in Pakistan).

Each participant will be asked to randomly select the first day of contact diary keeping from a set of 7 cards, each symbolizing a day of the week. One day before the selected first contact diary day, the participants will be trained in how to complete the diary. For adults or children unable to self-complete the diary, they will select a third-party person (henceforth called a “shadow”) to assist them in recording their contacts. Visit 3 will occurr one day after the second diary day. The field staff will assess the paper contact diary for completeness and inconsistencies and work with the participants to make any necessary amendments. Once the verification process is complete, data in the paper diary will then be keyed into the electronic diary by the field staff.

### Exit survey

The exit survey will capture additional details such as sentiments regarding the accuracy of the captured contacts, social and behavioral changes during the COVID-19 pandemic, and COVID-19 vaccination status.

### Wearable proximity sensors

We purchased an open-source off-the-shelf proximity sensor that was manufactured in the US, satisfies Federal Communications Commission (FCC) regulations, is relatively cheap, customizable, and small enough to be carried by people of all ages (figure 2). We selected the Adafruit Feather nRF52840 Sense sensor [https://learn.adafruit.com/adafruit-feather-sense] that uses active unidirectional Bluetooth Low Energy operating at 2.4 GHz to periodically transmit and receive data packets over short ranges relevant for respiratory infection spread.(25) It measures 51 mm x 23 mm x 7.2 mm and weighs approximately 6 grams. It has an on-board memory chip for data storage and is powered by a 3.7 V 500 mAh. We redesigned and updated the firmware used in the SocioPatterns project [www.sociopatterns.org] to be compatible with our new sensors. We also designed and 3D-printed tamper-proof, weather-proof plastic cases to enclose the sensor and battery. A fully assembled sensor with a battery and plastic case has a total cost of ∼$40 and weighs ∼20-30 grams. Prior to deployment, we conducted rigorous testing on case design, utility (ensuring radio communication between sensor dyads is achieved), signal propagation (how far signals are propagated, effects of multipath on signal strength), durability of battery charge and storage space capabilities (to establish optimum experiment duration).

**Figure 2.**
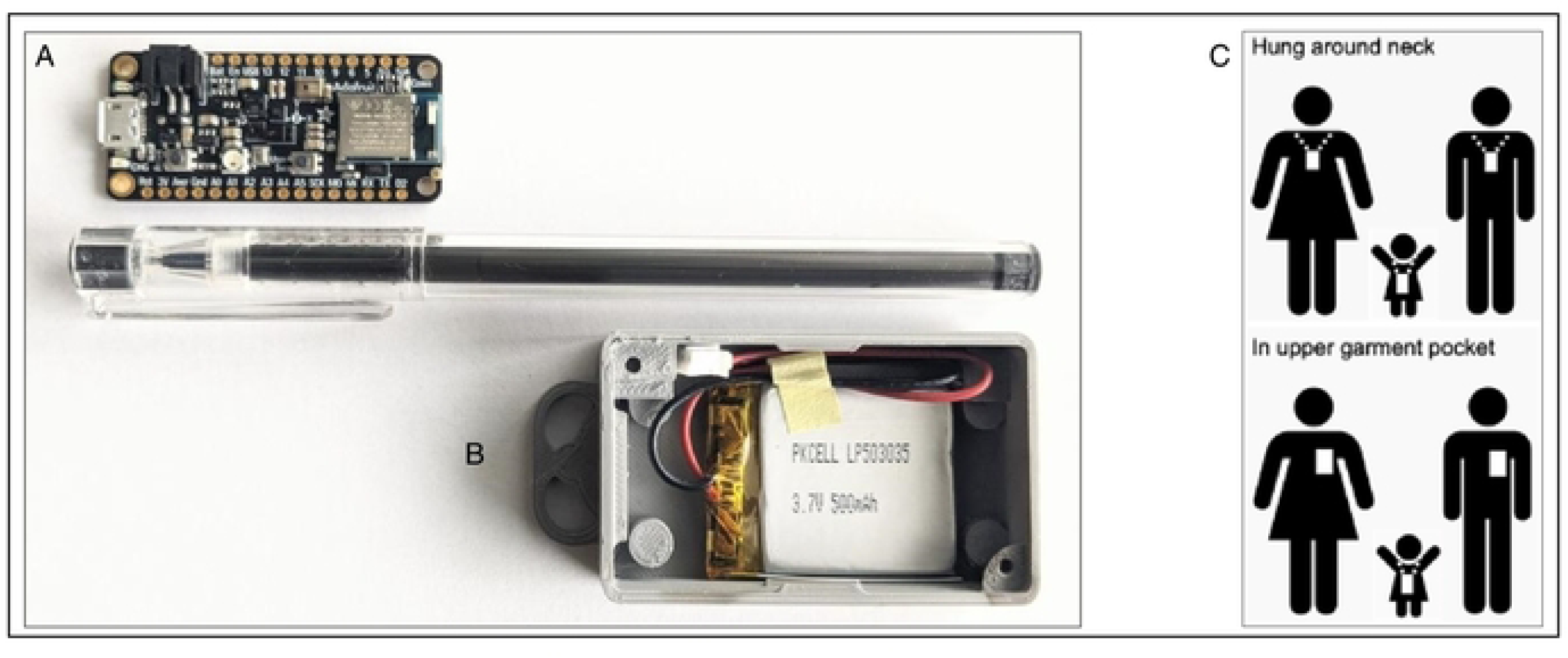
Schematic representation of the sensors. The sensor board consists of an antenna, flash storage device, and mini-USB port (A). A sensor will be powered by a battery and placed in a plastic case with a sealable weather-proof cover that will be disinfected in between each use (B). Sensors transmit signals only in the forward direction, thus only detecting close face-to-face interactions when hung around the neck or worn in a shirt/blouse pocket (C). The signals containing data packets are stored in the on-board memory of the sensor.

When in proximity, sensor dyads exchange data packets consisting of the sensor identifier (ID), timestamp, and transmission power level (received signal strength indicator, RSSI). Similar to previous studies, a ‘contact’ was defined as occurring between two individuals if their sensors exchanged at least one data packet over a 20-second interval. After a contact is established, its duration is defined as the interval in which the devices continue to exchange at least one packet for every subsequent 20s interval. Conversely, a contact is defined as broken if a 20-s interval lapses with no exchange of radio packets.(26) When worn on the chest area, signal exchange occurs when individuals are face-to-face within 1-1.5 meters of each other, thus defining proximity that may be relevant for transmission of respiratory infections. Assuming 8 hours of continuous data collection per day, the onboard sensor memory can log data packets for up to 7 days. It is stressed that the sensors detect face-to-face proximity between individuals by assessing the attenuation of the signals with distance but not an actual physical touch. Previous sensor deployment suggested that a 20-second interaction can be considered as a reasonable duration to describe “the fastest social interactions”, such as a quick conversation or handshake in a social gathering that may lead to the transmission of respiratory infections through droplets from coughing or sneezing.(26)

Once a battery has been inserted to activate the sensor, the sensor is sealed in a plastic case fastened with screws and additionally inserted into a cover that is disposed of after use to prevent potential infection transmission. We will request that the study participants hang the sensor at their chest by wearing it on a lanyard around the neck or insert it into the front pocket of a shirt/blouse or attach it to clothing using a clip. At the end of the study period, used sensors will be collected by study staff and the plastic covers will be disinfected. Data from each sensor will be downloaded onto a password-protected laptop. After data is downloaded, study staff will erase the data logs from the sensors and recharge them in readiness for the next deployment.

## Formative (Qualitative) Phase

The goal of the formative phase is to understand contextual factors and attributes of the community associated with the feasibility and acceptability of paper diaries and wearable proximity sensors in each site. This understanding will inform the design of data collection tools and implementation of the survey and other relevant study procedures.

### Selection criteria

Residents of the communities who consented to participate in the study **Error! Reference source not found.Error! Reference source not found.**were included in the qualitative study. Residents who **Error! Reference source not found.Error! Reference source not found.Error! Reference source not found.Error! Reference source not found.Error! Reference source not found.Error! Reference source not found.Error! Reference source not found.Error! Reference source not found.**did not consent to participate and non-permanent residents of the rural or urban municipalities **Error! Reference source not found.**were not included.

### Training of field staff

In-country study team members were trained in research ethics, data tools, and data collection methods before the initiation of study activities. Due to travel and congregation restrictions in place in the countries during the pademic, trainings were conducted online via Zoom, an online meeting platform. When restrictions were lifted, trainings were conducted in person. The training focused on the objectives of the formative study, recruiting methods, data collection tools, techniques of conducting focus group discussions, incorporating feedback on the FGD guide, data collection procedures, data management, translation, and transcription. Quality control of qualitative data and research ethics were also included in the training. After rapid analysis of the focus group discussions, cognitive interview guides were finalized, and the field staff were trained to conduct cognitive interviews (CI). The data were analysed on-site.

### Focus group discussions

A focus group discussion guide designed with the assistance of the team anthropologist was used to facilitate the discussions. The questions in the guide focused on identifying community perceptions and assessing the acceptability and feasibility of our study procedures, and the use of the contact diary and sensor. Participants also discussed potential barriers and suggested solutions to overcome any challenges within their families or communities. Thirteen focus group discussions were conducted per study site. The following groups were recruited to participate in the FGDs: 1) community and religious leaders, 2) parents/caregivers of infants < 6 months old, 3) male and 4) female community or household elders, and 5) teenagers.

#### Sample size

Between 1-2 FGD activities were conducted per site for each group, with 6-8 participants per activity. To counter refusals and no-shows, we invited eight to twelve participants to each FGD. However, we maintained a minimum of 6 participants per discussion group. The sessions were held at a location convenient to participants in the community and conducted in a local language chosen by participants, after obtaining their consent. During the FGDs, participants received an explanation and demonstration of the use and operation of the proximity sensor and paper contact diaries through illustrative images and simulations of use and function. Participants provided their feedback on the design, language, ease of understanding, sociocultural barriers, and the study implementation process and use of these tools. Each FGD lasted 60 minutes. All FGDs were recorded and translated to English for analysis.

### Cognitive interviews

During cognitive interviews, we clarified through a few carefully selected questions on how community members understand and interpret survey questions and procedures, specifically what these questions mean in the local context. The interviews helped to ensure that survey items achieved their measurement purpose and elicited the intended meaning of the question. With the assistance of the team anthropologist, we developed a cognitive interview guide that included select survey and contact diary questions that were difficult to understand during FGDs. The cognitive interviews identified comprehension and language problems, i.e., not knowing the meaning of words/phrases, or having different local or cultural terms that are more appropriate or better understood; inclusion/exclusion problems, i.e., determining whether certain concepts are to be considered within the scope of an item, and whether caregivers include or exclude experiences within the intended scope of the question; temporal problems, i.e., the period to which the question applies; logical problems, e.g., how participants interpret certain phrases in survey questions, and computational problems, e.g., readability of items, and guessing age of contacts. The interviewer starts by obtaining written consent from participants. Participants were then asked to read each question, and before responding with an answer, they paraphrased the question in their own words for the interviewer to ensure that they understood the meaning correctly. While answering the question, they could also ‘think aloud’ to share what was going through their mind when they are considering the response options. Participants were then asked to complete a template of actionable recommendations based on the interviews. A separate note-taker helped to document the interview. The cognitive interviews took place with 3 individuals per age group per site. Six age groups were covered for at least 18 individuals in each study site.

### Recruitment procedures

Appropriate strategies were used to recruit participants to each site. First, we held meetings with local relevant stakeholders such as local authorities (chiefs of administrative posts and localities, heads of the villages, chiefs of neighborhoods or any other authority), religious leaders, and health practitioners to explain the purpose of conducting this study and identify potential participants.

Meetings were held according to the epidemiological situation and the COVID-19 alert system and guidelines from the Ministry of Health. A strict protocol of facial masking, hand hygiene, social distancing, and a well-ventilated room were followed. The dates and venues of meetings were defined according to the availability of the participants and according to the epidemiological situation and COVID-19 alert system of the Ministry of Health for the various sites. Where demographic surveillance data were available, participants were selected at random based on 10 age groups identified for the quantitative phase (i.e., <6 months, 6-11 months, 1-4 years, 5-9 years, 10-14 years, 15-19 years, 20-29 years, 30-39 years, 40-59 years, and 60+ years). In addition, we used the following methods of recruitment for specific groups:

i. Community and religious leaders were identified with the support of the heads of localities and of the administrative post, and religious leaders’ group with support of the religious affairs commission, where large parts of the religions that were present in the study area are represented.
ii. Elders were recruited with the support of neighborhood chiefs and community leaders based on selection criteria discussed with the study team.
iii. Parents of infants were identified by generating a list of all infants aged up to 6 months of age as of data collection time, and identifying the names of their parents and household locations.
iv. Teachers were identified with the support of the principals or pedagogical heads of schools from the study area, based on criteria to be discussed with the study team.

**Table 2.** Recruitment period for qualitative phase.

| Study Sites | Recruitment Start Date | Recruitment End Date |
| --- | --- | --- |
| Mozambique | 04 August 2020 | 16 October 2020 |
| Guatemala | 19 July 2021 | 05 November 2021 |
| India | 16 February 2022 | 12 April 2022 |
| Pakistan | 14 Dec 2022 | 18 Feb 2023 |

### Qualitative data analysis

Qualitative data were analyzed and suitable recommendations for updating the survey questions were finalized with input from the principal investigators. Following each focus group discussion meeting, the facilitator prepared a summary of impressions. The notes and audio recording from the focus group discussion were transcribed and translated into English and study team members reviewed the transcripts. Rapid analysis techniques (27,28) were applied to achieve results informing the main study tools. Separate coding frameworks were developed for focus group discussions and cognitive interview data. The thematic analysis method (29,30) was used to analyse the transcripts and identify and organize themes into clusters and categories through coding. The analysis included reading the extended transcript summaries several times to achieve an understanding of the meanings conveyed, identifying important phrases, and articulating and confirming meanings through research team discussions to reach agreement. Respondents’ answers to each question were grouped, organized, and classified into categories. Information obtained informed the development and refinement of the quantitative phase tools and procedures for each respective site. Recommendations were made for modifications to the quantitative phase study procedures and tools as appropriate.

### Qualitative data management

At each study site, coinvestigators supervised the process of data collection. All audios, interviews, and summaries were coded and saved in the local team’s servers. All data will be shared with Yale University and Emory University team members through a secure online data sharing and storage platform. Only trained team members will be able to access the documents on this platform. At all sites, all focus group discussions and cognitive interviews were audio-recorded, and all texts or audio captured were transcribed in preferred local languages and then translated into English verbatim. The languages used at each site were as follows: Portuguese in Mozambique; Spanish in Guatemala; Tamil in India, and Urdu and Sindhi in Pakistan. Transcripts and field notes were anonymized through the redaction of identifying details prior to translation. Digital recordings of FGDs and cognitive interviews were destroyed after transcription; therefore, only the researchers conducting the FGDs, and cognitive interviews and the transcriber had access to unredacted data. All quotations or descriptions used in reports were anonymized, and additional potentially identifying details will be redacted or altered to further protect anonymity. All photographs and video footage were only taken with consent and would be appropriately anonymized. To ensure privacy, participants were requested to choose between the actual name or nickname for the contact person or location. All signed consent forms were stored by the data manager in a locked cabinet in the office.

## Quantitative Phase

The goal of the quantitative phase is to collect and summarize data on the rates and characteristics of social contacts using social contact diaries and electronic proximity sensors. Quantitative data collection was scheduled in phases across the sites over two years.

**Table 3.** Recruitment period for quantitative phase

| Study Site | Recruitment Start Date | Recruitment End Date |
| --- | --- | --- |
| Mozambique | 05 April 2021 | 05 April 2022 |
| Guatemala | 03 March 2022 | 22 December 2022 |
| India | 25 July 2022 | 04 February 2023 |
| Pakistan | 02 June 2023 | Ongoing |

## Contact diary study

### Sample size

Our study sample will be divided into 10 age groups (<6 months, 6-11 months, 1-4 years, 5-9 years, 10-14 years, 15-19 years, 20-29 years, 30-39 years, 40-59 years, and 60+ years). We framed our sample size calculations to detect sufficient precision in infants <6 months old, an age group of high interest. We powered each age group to have a standard deviation consistent with the precision in the estimate of the youngest age group (0-4 years) in the POLYMOD study (standard deviation = 7.65 contacts per day) (31). We chose to base our sample size calculation on the youngest age group because infants less than six months of age represent the smallest fraction of the overall population size and whose data are largely unavailable since they require a literate older person to keep records on their behalf (19). We inflated the sample size by 10% for each age group to account for attrition due to study withdrawal and loss to follow-up. This resulted in a sample size of 63 per age group per site, thus 630 for each site, 1260 per country, and 5040 participants overall.

## Electronic proximity sensor study

### Sample size

We will recruit 63 households per site (126 per country). At each site, households with a child aged <6 months, 6-11 months, 1–4-years were selected randomly and proportional to these age groups.

### Participant selection

Once we locate a household, we will make an initial visit (visit 1) to seek permission from the head of the household to allow us to engage the other household members. In Guatemala only, where we do not have sampling frame, we identified household clusters around a 200 m^2^ radius using satellite imagery from Bing. From a central location in each cluster, we approached all households in sequence until we attained our target sample size. We will obtain written informed consent (adults ≥18 years old) or assent (for children aged <18 years).

### Study procedures

To train all the participants in diary-keeping and sensor-wearing procedures, we will request the head of the household to select a day of the week when all household members will be present at the same time. On this selected day (start day), we will visit the household and train the members on the diary and sensor study procedures (figures 3 and 4). Each participant will wear the sensor over 7 continuous days for data collection. Participants will be required to doff the sensor when not in use such as when sleeping or leaving the house, or when encountering water. In addition, the head of the household will select one day at random (diary day one) between the start day and end day (7 days after start day).

**Figure 3.**
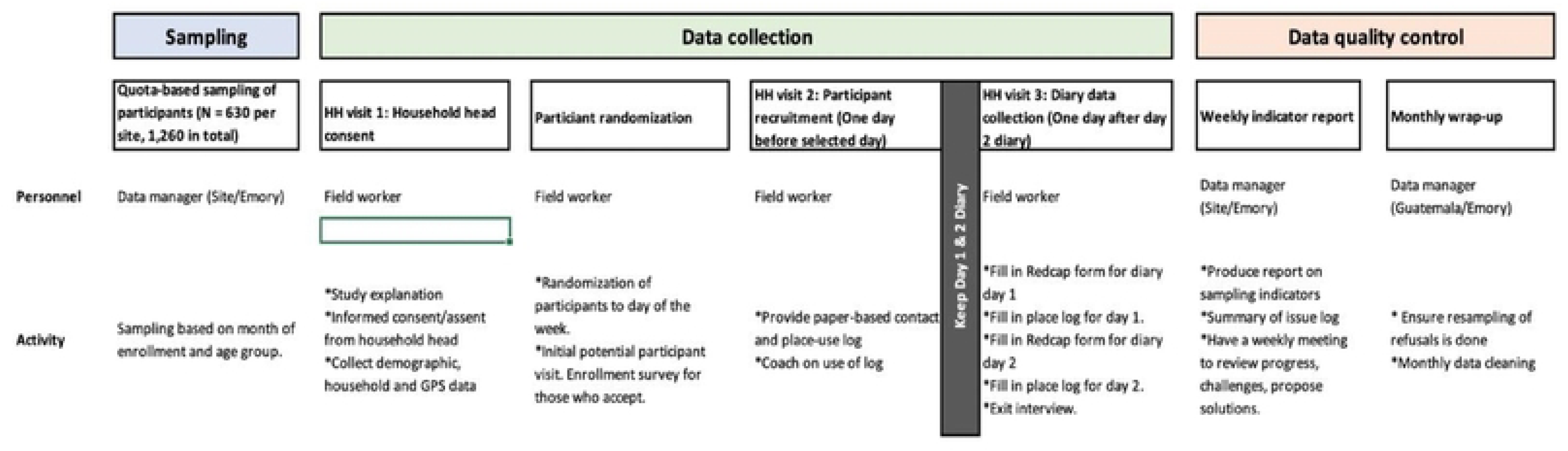
Flow of field visits showing data collection procedures for the three visits (contact diary study)

**Figure 4.**
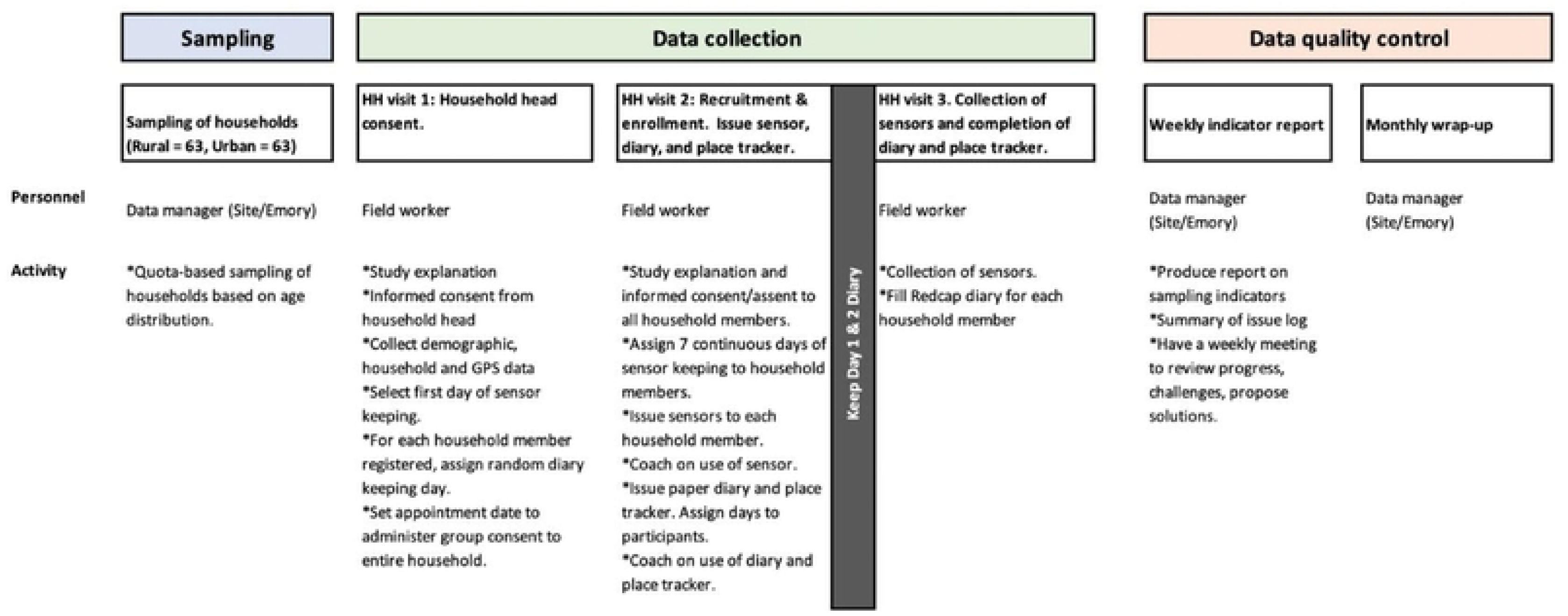
Flow of field visit showing data collection procedures for the three visits for the (proximity sensors study)

Each household member will be required to keep a record of their contacts in the paper diary on two consecutive days. The research team will record the date and time when the sensors were issued and collected, while participants will keep a daily log of when the sensors were worn or taken off. Data from each sensor will be individually downloaded into a laptop at the site office.

## Quantitative data analysis

### Construction of age-specific contact matrices

The primary outcome from this analysis will be age-specific contact matrices containing the average (or expected) number of contacts made per day between age groups combinations. We will create three age-specific contact matrices for each study site: a matrix of physical contacts only, a matrix of nonphysical contacts, and a matrix of all contacts (both physical and nonphysical contacts).

### Identifying predictors of number and duration of contacts

We will investigate whether certain demographic and biologic characteristics are predictors of the frequency and nature of contacts. This regression analysis will result in the identification of key drivers of contact.

### Defining contacts using wearable sensors

To comprehensively profile the frequency, duration and intensity of social contacts of young infants with their household members, we will analyze high resolution data collected using wearable proximity sensing devices. Data collected from each participant will be obtained and cleaned separately. In sensitivity analyses, different time-interval definitions of a ‘contact’ will be considered.

### Validation of diary-based collection (Sensor Study)

We will use the sensor data to validate the diary data using two methods. First, using variables that characterize each contact such as age, sex, duration, and relationship, we will identify contacts that were identified by [1] diary, [2] sensor, or [3] both. We will then calculate the level of agreement between diary and sensor data using Cohen’s kappa coefficient (k). Second, using sensor data, we will construct age-specific contact matrices containing the average number of contacts made per day using the methods described in the contact diary study. We will compare these matrices with those generated from diary data but restricted to household contacts using the coefficient of accuracy (C(a) and the coefficient of determination (r_c_). C(a) measures the accuracy and r_c_ measures the precision of the relationship between the two data collection methods (32). We will then perform the same comparisons for data on the duration of contacts.

### Characterizing the social contact of young infants

We will generate a detailed characterization of the social contacts of infants aged <6 months with their household members. To do this, we will calculate the infant proximity score for each household member, which is defined as the fraction of time spent by each household member in proximity with the infant, over the total infant contact time.(33) Certain demographic and behavior characteristics (e.g., maternal age, sex, household size, and country) are predictors of the infant proximity score. To evaluate the association between social contacts and these predictors, we will use regression methods such as negative binomial regression where the dependent variable is the infant proximity score.

### Estimating effects on total number and duration of contacts

We hypothesize that certain demographic and behavior characteristics (e.g., maternal age, sex, household size, and country) are predictors of the frequency and nature of social contacts of young infants. Identifying these factors will aid in generalizing our findings and serve as a guide for other researchers to select the right contact matrices and determine what predictors are important to include in future analyses. To evaluate the association between predictors and the total number or duration of contacts, we will use similar regression methods as above. In addition to providing an analytical understanding of the determinants of contact frequency and types, the results from this analysis will help to guide researchers on which contact matrix is best suited to a given modeling application or generalizable to similar populations.

### Creation and analysis of an aggregated contact network

We will create an undirected social contact network at the household level, aggregated over the whole study period per site, draw network graphs, and calculate descriptive network statistics. Nodes represent study participants and edges are the presence of at least one recorded contact event between individuals during a contact window (figure 5.) Edge attributes will reflect the type or intensity of the contact (i.e., cumulative duration of contact). The attributes of the nodes will be included using the demographic data and relationship to participant as collected during the initial interview of study participants. We will calculate [1] the degrees of each node (k) and describe the distribution by degrees P(k); [2] the distance (d) between nodes (e.g., the shortest distance between individuals in households) and the average distance𝑑 (𝑑); and [3] the intermediation centrality defined as the proportion of the shortest paths that pass through a single node Bi. These statistics can subsequently be used to explicitly model disease transmission in networks.[27] We will also use an adjacency matrix (A*_ij_*) to summarize connections made with our populations. Using the adjacency matrix, we will calculate the average number of contacts according to the following formula: 𝑛 = _𝑁_𝛴_𝑖𝑗_𝐴_𝑖𝑗_, where N is the population size (i.e., size of the households).(35)

**Figure 5.**
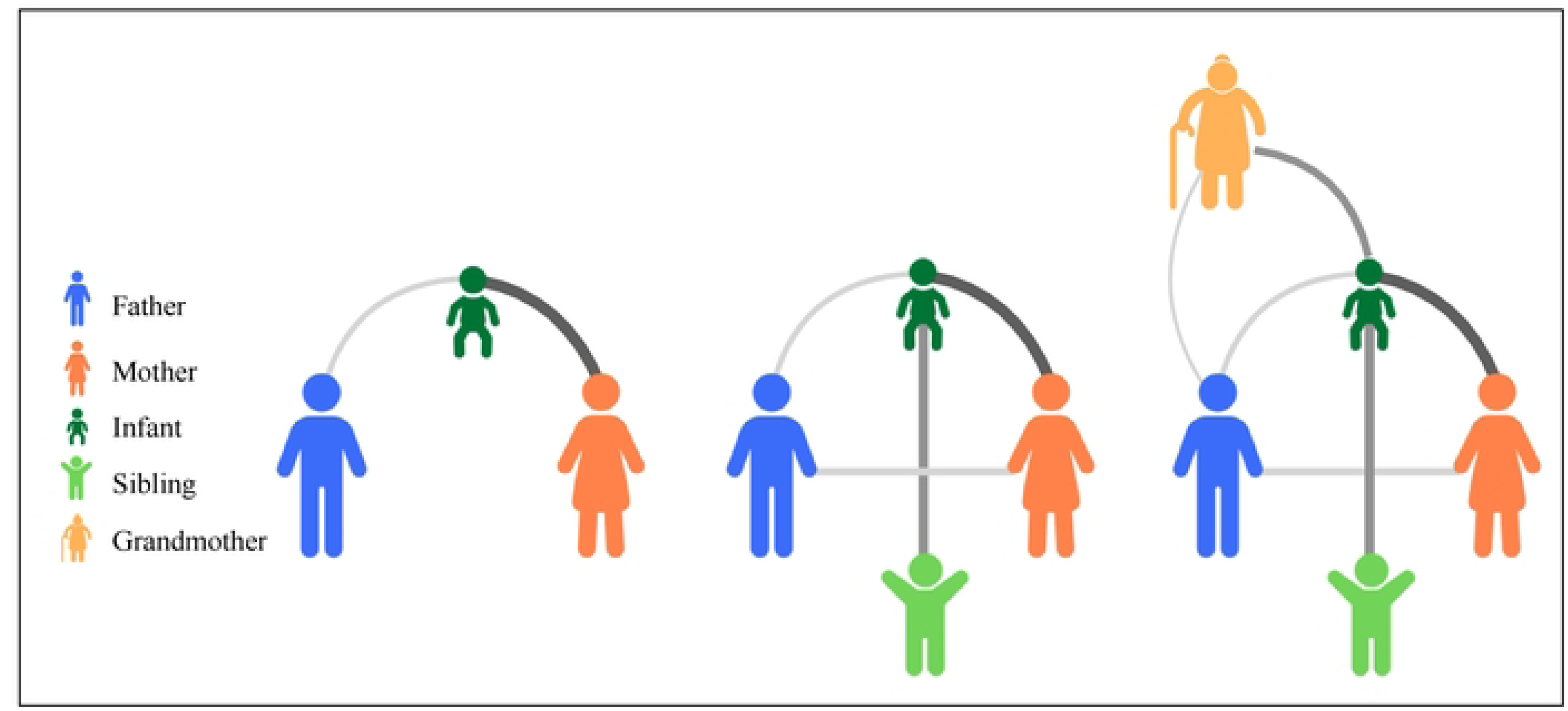
Household contact network. Contact networks will be visualized in order to depict the presence, type and characteristics of contacts (edges). Here, as an example, household members are represented as nodes color coded for relationship to the infant. The thickness of the edges represents cumulative time in contact, with thicker/darker edges representing more time spent in contact.

## Quantitative data management

Study data will be collected and managed using REDCap electronic data capture tools hosted at Emory University(36,37). REDCap (Research Electronic Data Capture) is a secure, web-based software platform designed to support data capture for research studies, providing 1) an intuitive interface for validated data capture; 2) audit trails for tracking data manipulation and export procedures; 3) automated export procedures for seamless data downloads to common statistical packages; and 4) procedures for data integration and interoperability with external sources. A different project will be created for each country, such that country study teams will only have access to country-specific data.

### Security of the sensors

Signals in the power range emitted by the sensors do not pose reported radiation harm while in use by humans. Additionally, the radio system for these sensors is approved for use by the United States Federal Communications Commission (US FCC). The sensor is FCC certified and licensed under 47 CFR part 2, which means that it does not cause harmful interference to radio signals from household appliances. Additionally, the beacon is certified to comply with rules limiting other policy objectives - such as human exposure limits to radio frequencies and hearing aid compatibility (HAC) with mobile wireless devices. The sensor and battery will be stored in a small weather resistant box that is sealed with two small screws. This box will then be inserted into an additional outer protector that will be discarded after each use. A strap will be attached to the outer protector, so that the participant can use it hanging around the neck, and it will be adjusted so that it is kept at chest level. For use in young children, an appropriate hook will be provided so that it can be attached to the participant’s clothing. Parents will be shown how to attach the sensor onto children’s outer garments.

### Data security specifications

The data exchanged between the sensors is automatically encrypted during its internal storage. We have developed programs using Python software that data managers will run to automatically download data from sensors. The sensors must be connected to the computer using data cables before the data can be downloaded. Only data administrators will have access to these programs. The decryption of the data will be done by the data analyst.

## Ethics approval and consent to participate

The study protocol and all qualitative and quantitative survey tools were submitted to the Institutional Review Boards (IRBs) of all the study sites as well as the national ethics committees of all the international sites before initiation of study activities. This study addresses household and individual information and all data collection shall follow institutional, national, and international policies and procedures or guidelines for protecting the information provided by participants and maintaining the confidentiality of participants and their contacts. To comply with this, the field staff were trained in the need to ensure that data collection is done accordingly and ensure maximum confidentiality possible under field conditions. Informed consent to participate was taken from parents/legal guardians of minor participants. For all the sites, we developed the following consent and assent forms for both qualitative and quantitative data collection activities: Consent for Adult Participants (applicable to individuals aged 18 years or older to give consent for themselves to participate in the study); Consent form for Parents and Guardians of Individuals under 18 years old (applicable to parents and legal guardians of children under the age of 18 years to give consent for them to participate in the study); Assent form for Adolescents aged 12 to 17 years (applicable to adolescents aged 12 to 17 years old to give assent for themselves to participate in the study); and Consent for Household Heads (applicable to heads of households to give consent for themselves and all members of their household to participate in the study). In Guatemala, we obtained either written or verbal assent for children older than 7 years for focal group discussion and cognitive interviews, upon request of the community. This was approved by the UVG IRB. Ethical approval of the study was obtained from the Institutional Review Board for all the sites as well as at Emory University before the onset of study activities. Ethical approval dates and numbers for all study sites:

| Site | Emory Approval date | Approval number | Site approval date |
| --- | --- | --- | --- |
| Parent (Emory University) protocol | March 4, 2020 | 00105630 | 3/04/2020 |
| Yale University Protocol | **Reliance Agreement:**<br>November 11, 2019 | 2000026911 | 6/12/2019 (Yale IRB) |
| Mozambique | August 9, 2020 | CIBS-CISM/012/2021 | 3/26/2020 (CISM) |
| Guatemala | July 21, 2021 | 223-12-2020 | 1/25/2021 (National IRB)<br>3/03/2021 (UVG IRB) |
| India | June 26, 2019 | IRB Min. No. 12065 | 2/02/2022 (CMC IRB) |
| Pakistan | December 22, 2022 | 2022-7912-23648 | 12/22/2022 (AKU ERC) |

## Status and timeline of the study

Qualitative data collection has been completed in three sites and is ongoing in the fourth site. IRB approval has been obtained in all the sites. Quantitative data collection is expected to be complete by March 2024.

## Discussion

This multicountry cross-sectional study using qualitative (formative phase) and quantitative data from Mozambique, Guatemala, India, and Pakistan is the first study to comprehensively characterize the patterns of social contact and mixing in urban and rural LMIC settings using standardized social contact diaries. We will also profile the social contacts of infants with their household members using contact diaries and wearable proximity sensors. The deidentified data collected in this study will be made publicly available for use in dynamic transmission models of infectious diseases. These empirical data will allow a more accurate representation of human interactions that lead to the transmission of pathogens transmitted through close contact.

The qualitative, formative phase of this study is unique and will generate useful information regarding perceptions and acceptability of contact diaries and wearable proximity sensors to collect data on social contact and mixing patterns between individuals in LMICs. This information will be used for developing more complex studies that use contact diaries and wearable proximity sensors. Finally, conducting this study in LMICs will generate results that may be more representative of contact patterns in such setting. Settings. Currently, many modelers use European data from POLYMOD or POLYMOD data projected onto LMIC populations.

Despite the use of rigorous mixed qualitative and quantitative methods, we anticipate some challenges. Participants may only recall contacts that are recent or salient (those that are most important, such as meeting family or friends compared to strangers). This may lead to an underestimation of contact rates. We expect that children who are not in school and those aged less than 10 years old may not be able to self-report their contacts. Additionally, adults who are unable to or have limited ability to read and write may not be able to record their contacts in the paper contact diaries. For these groups, we will assign parents/guardians as “shadows” to the children to help with recording contacts. Then adult participants will be asked to select an individual to record their contacts on their behalf. This would require adequate training of both participants and shadows to understand diary keeping procedures while respecting the privacy of the participants. Furthermore, to profile the social contacts of infants with their household members using contact diaries and wearable proximity sensors, only interactions between participants wearing sensors will be recorded. Sensors do not record touch or other characteristics of interactions between participants. Therefore, while the sensors provide accurate and time-resolved data on physical proximity, they lack the richness of the characteristics of the contact. The site-specific contact matrices and all underlying data will be made publicly available. Study findings will be published in scientific journals and presented at national and international scientific and programmatic forums.

## Data Availability

All data and code generated during the current study will be made available via our GitHub repository. The data will also be made available to download on Zenodo.

## Authors’ contributions

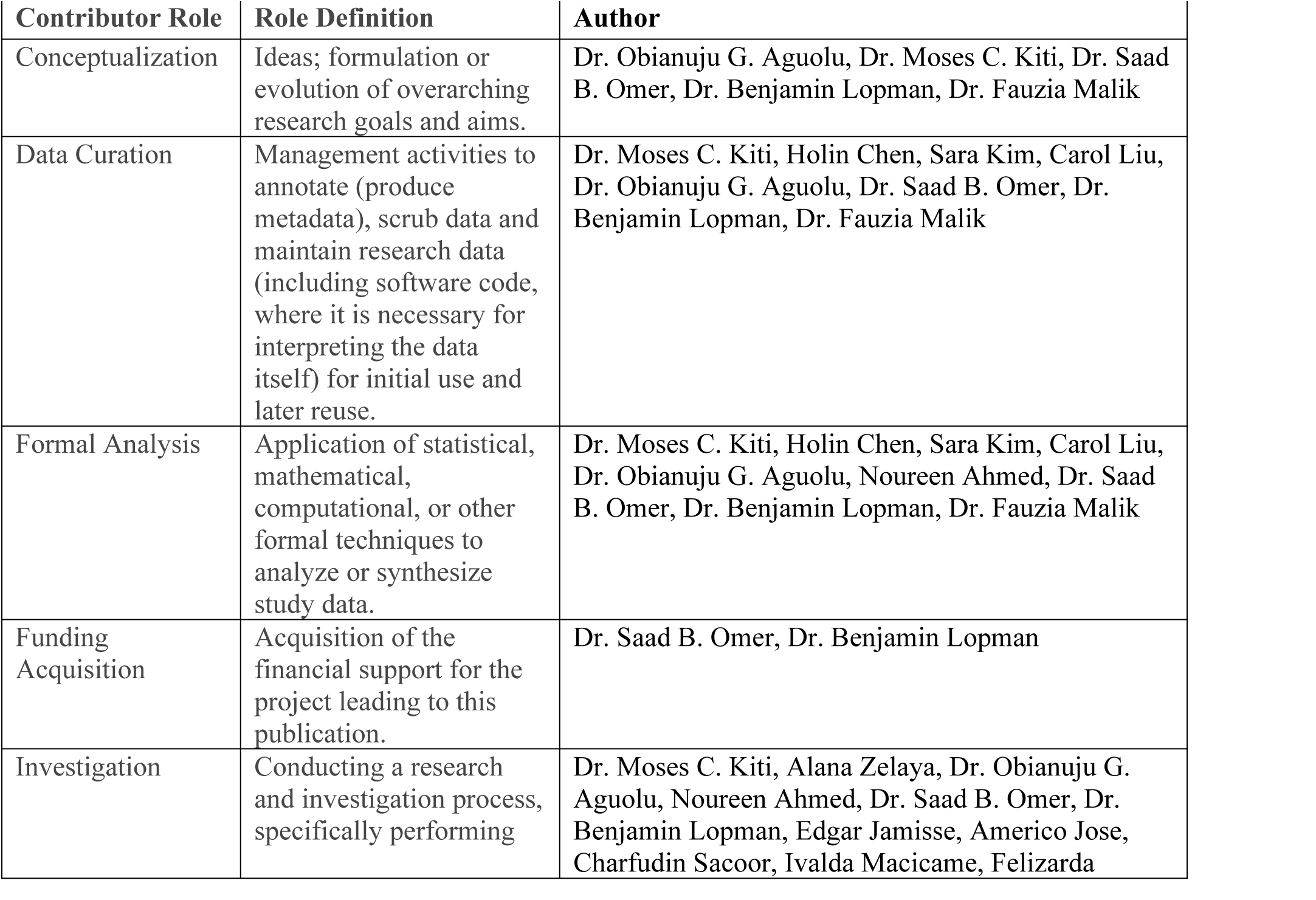
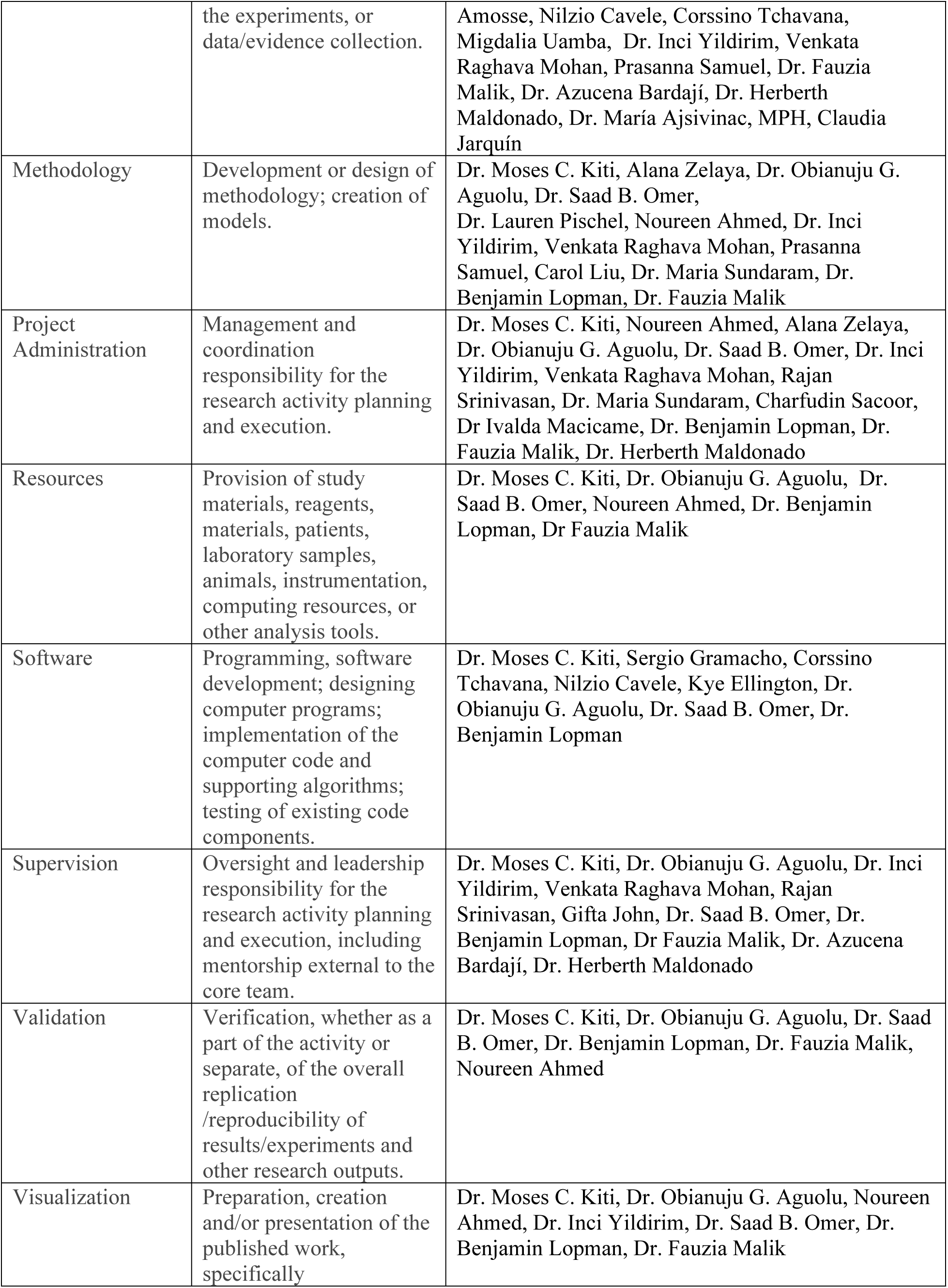
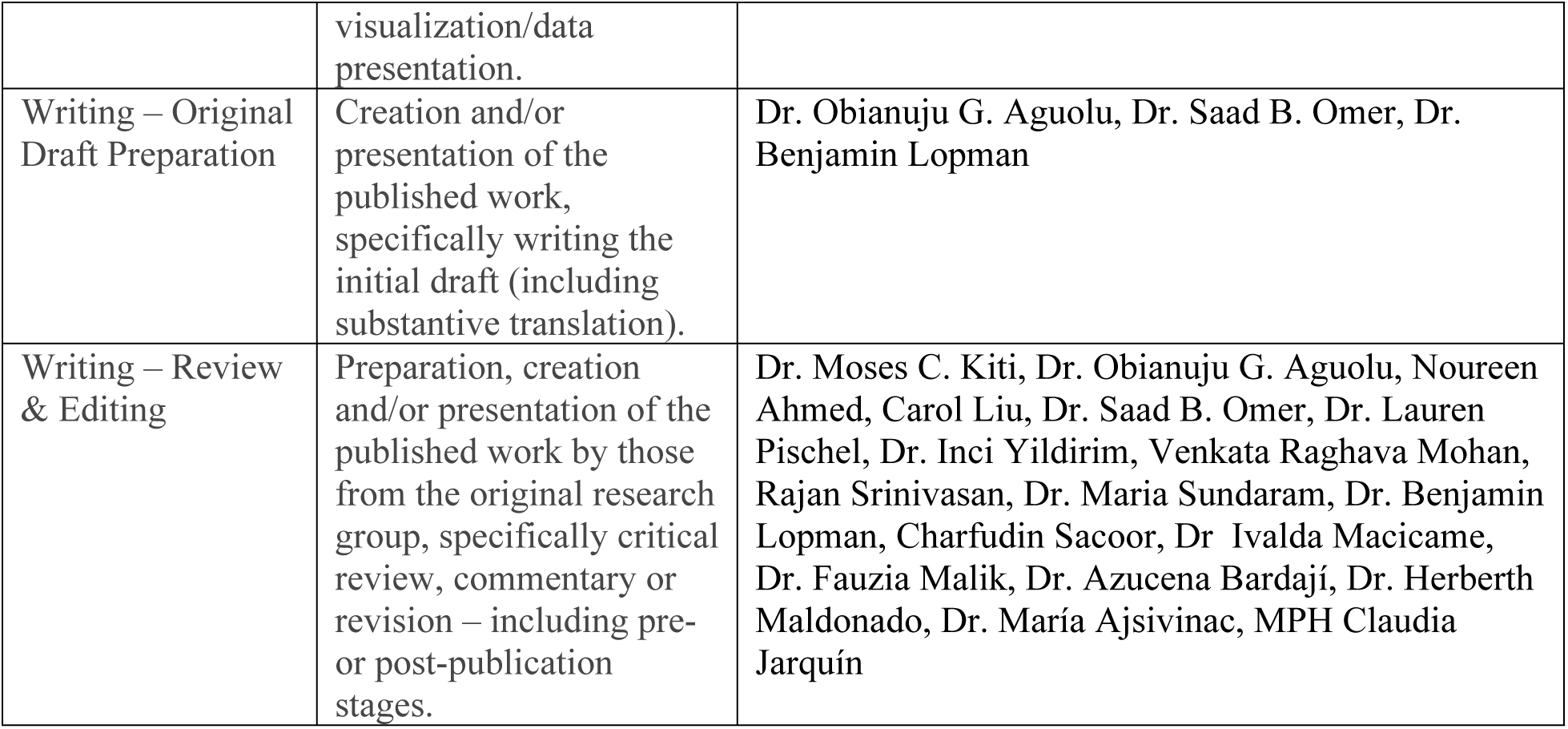

## Acknowledgement

We thank the many contributors to this study for their expertise and assistance throughout all aspects of our study and for their help in writing the manuscript.

## Supporting information

1. Survey:

a. Enrollment survey
b. Electronic contact diary
c. Exit survey
2. Paper contact diary
3. Places diary
4. Focus group discussion guide
5. Cognitive interview guide

## Figures

1. Figure 1. Global map showing the location of Guatemala, India, Mozambique, and Pakistan.
2. Figure 2. Schematic representation of the sensors.
3. Figure 3. Flow of field visits showing data collection procedures for the three visits (contact diary study)
4. Figure 4. Flow of field visit showing data collection procedures for the three visits for the (proximity sensor device study)
5. Figure 5. Household contact network.

## List of abbreviations

LMICs: low-and-middle-income countries
FGDs: focus group discussions

